# Community Collaboration in Public Health Genetic Literacy: Co-Designing Educational Resources for Equitable Genomics Research and Practice

**DOI:** 10.1101/2024.05.24.24307892

**Authors:** Juhi Salunke, Grace Byfield, Sabrina N. Powell, Daniel Torres, Grace Leon-Lozano, Jahnelle Jackson, Andreas K. Orphanides, Jonathan Shaw, Thomas Owens, Jonathan S. Berg, Elizabeth Branch, Lennin Caro, Stefanija Giric, Julianne M. O’Daniel, Bradford C. Powell, Ken Ray, Megan C. Roberts, Carla Robinson, Samantha Schilling, Nicole Shaw, Erin Song, Margaret Waltz, Ann Katherine M. Foreman, Kimberly Foss, Laura V. Milko

## Abstract

**Introduction:** Unequal representation in genetic and genomic research is due to various factors including historically inequitable and unjust institutional research practices, potential mistrust of biomedical research among underrepresented populations, and lack of access to or awareness of research opportunities. Facilitating sustainable dialogue between diverse communities and genetic researchers can cultivate trusting, bidirectional relationships, potentially encouraging greater participation in research. Herein, we describe the co-creation of public health educational materials and dissemination plans.

**Methods:** We describe co-creation by genetics clinicians, researchers, and community members of Genetics and Genomics Educational modules. These modules are aimed at enhancing genetic literacy with a goal to facilitate informed decision-making regarding genetic research and health services. We used Designing for Dissemination and Sustainability, which is grounded in Dissemination and Implementation science, and the Fit to Context process framework to guide the process. This approach ensures the public health context and diverse audience for the modules are considered throughout their development. Additionally, it ensures that broader goals such as dissemination, equity, and sustainability are integrated from the outset, fostering long-term impact and effectiveness.

**Conclusion:** This article offers an evidence-based template for adoption or adaptation by other community-engaged groups, aimed at bolstering equity and sustainability in the development of health care interventions, with an emphasis on accessible public health literacy. The co-creation of both materials and dissemination plans between researchers and community members may improve the cultural appropriateness and relevance of public health genetics campaigns. Ongoing research is needed to assess the impact on receptiveness and participation.

## Introduction

Rapid progress in DNA-based population screening, focusing on actionable disease-predisposing genetic variation in asymptomatic individuals, holds tremendous potential to mitigate adverse health outcomes and drive the advancement of precision public health [1]. However, widespread application of this technology in a healthy population faces numerous challenges to acceptance by the public, exacerbated by insufficient research in communities that reflect the population at large [2,3]. Research on participation in clinical translational genomic studies reveals a disproportionate representation of higher-resourced individuals and those with European ancestry, while lower-resourced and racially minoritized populations may be apprehensive about the disclosure of genetic data, its security, potential misuse, and the associated costs and risks of future discrimination [4–8]. Due to a lack of trust in and knowledge about genetic and genomic research, if only specific segments of society engage in genomic research existing health disparities will be exacerbated in clinical practice [9–11].

Mistrust of and misinformation about genomics, particularly in historically marginalized groups, must be addressed before genomic technology can be equitably applied to improve public health outcomes. Numerous studies have found that multilingual and culturally responsive educational resources could build genetic health literacy and awareness, and subsequently improve participation in research [12–15]. Research has shown the profound impact of fostering trust-based relationships and bidirectional communication, emphasizing the inclusion of communities historically marginalized and exploited by biomedical research, notably Black and African American people who have been systematically excluded from the advances made with their contributions [16–21]. This inclusive approach has been shown to enhance interest and participation in research, particularly when intended beneficiaries are engaged throughout the development process [22–26].

However, most publicly available genetic educational resources are tailored towards individuals with higher education levels or a background in science or medicine, often overlooking underserved communities with lower literacy levels. These materials typically concentrate on disease-specific or test-related information, which might not effectively convey essential genetic concepts to a broader audience [27,28]. Furthermore, educational materials aimed at patients frequently lack input from the very communities they seek to serve, resulting in limited relevance and effectiveness [29,30], creating a gap between the evidence-based interventions’ design and their adoption by the communities for which they are intended.

The Age Based Genomic Screening (ABGS) study, funded by the National Human Genome Research Institute and led by researchers at the University of North Carolina at Chapel Hill (UNC), seeks to integrate targeted genomic screening into routine pediatric well-child visits. At the core of the design and development of the ABGS study is a robust commitment to community partnership throughout its duration, along with the application of models and framework from Dissemination and Implementation (D&I) science with the goal of enhancing access and equity across diverse settings[31].

One such model, Designing for Dissemination and Sustainability (D4DS), encompasses four key phases during the development of a new health intervention —Conceptualization, Design, Dissemination, and Impact—aimed at ensuring the fit of an intervention with the contextual characteristics of its target audience and setting, and planning for active dissemination and sustainability from the outset of development [32]. Throughout the development of an intervention, process, determinants, and evaluation frameworks from D&I science provide structure for the D4DS methodology. The Fit to Context (F2C) process framework underpins and organizes the D4DS elements to enhance success across each of the four phases. Determinants frameworks, such as the Practical, Robust Implementation and Sustainability Model (PRISM)[33], prioritize understanding of beneficiaries, adopters, and contextual characteristics, ensuring iterative alignment between the intervention and its evolving context. Outcomes frameworks such as Reach, Effectiveness, Adoption, Implementation, and Maintenance (RE-AIM) [34], facilitate iterative evaluation ensuring continued alignment with context and equitable reach, adoption, sustainment, and health impact over time [35]. This thorough approach optimizes feasibility and adoption among diverse audiences through ongoing community partnerships, feedback, and evaluation, thereby fostering health equity through tailored strategies and systemic changes.

ABGS is a novel proposal aimed at expanding screening opportunities for children for actionable genetic conditions, building upon the established public health success of newborn screening (NBS), which universally screens newborns for conditions offering clear health benefits through early detection and treatment. Unlike NBS, which operates on an ‘opt-out’ basis due to its critical role in public health, ABGS seeks to expand the scope of screenable conditions, including some with lower penetrance and later onset in childhood. This expansion necessitates a shift to an ‘opt-in’ approach. The transition to “opt-in” screening for ABGS requires the establishment of new mechanisms to guide parents and providers through the complex decision-making and consent processes inherent in the program. Recognizing this need, we aim to bridge this gap by creating educational resources on public health genomics developed through participatory design with community members. Our approach is rooted in the D4DS foundation, operationalized by the Fit-to-Context (F2C) process framework, as we outline our current and future goals for development.

## Methods

### Application of the Fit-to-Context (F2C) Process Framework

We use the F2C framework, building upon the foundational D4DS principles and methods, to guide the development of resources aimed at enhancing public health genetic literacy and fostering equitable participation in genetic and genomic research. Expanding on D4DS, the F2C framework pairs actions and outcomes for each of the iterative phases of Conceptualization, Design, Dissemination, and Impact with specifically relevant research methods and approaches. The adoption of the F2C facilitated the systematic application of D4DS approaches, drawing from diverse fields such as D&I, communications, marketing, and the arts. The goal is to craft a public health intervention tailored to its target audience, with considerations for future dissemination and sustainability from the project’s inception (Figure 1).

**Figure 1:**
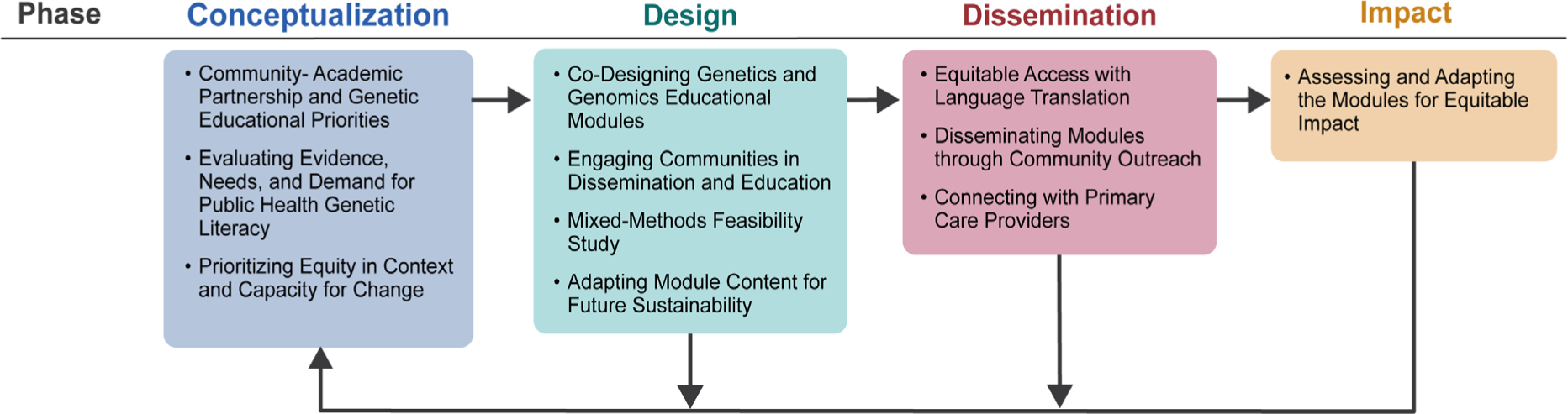
Designing for Dissemination and Sustainability (D4DS) phases, with adapted constructs based on the Fit to Context (F2C) process framework, used in the development of the Genetic and Genomic Educational Modules. The application of F2C guides the process, ensuring contextual alignment to meet diverse educational needs while advancing dissemination, equity, and long-term sustainability objectives.

### Phase 1: Conceptualization

The initial Conceptualization phase of D4DS underscores the importance of community partnerships in understanding the need, demand, and capacity for a new intervention aimed at addressing a health issue of significance to communities. The subsequent subsections outline our prioritization of the F2C objectives and outcomes for the Conceptualization phase in developing public health genetics educational resources. These priorities include establishing a community-academic partnership, consulting primary literature to gain insight into known and established community needs and demands for such resources, as well as addressing contextual factors essential for health equity and capacity for change.

#### Establishing Community-Academic Partnership and Genetic Educational Priorities

In 2021, UNC researchers assembled a Community Research Board (CRB) comprising parents from diverse backgrounds across NC and involved them as active research collaborators [23]. Currently, the approximately 25 person (membership varies slightly due to occasional attrition and new recruitment) ABGS collaborative research team comprises a multi-disciplinary team of researchers and clinicians from UNC with expertise in relevant fields (including bioethics, social science, education, community engagement, newborn screening, dissemination and implementation science, health communication and decision-making, genetic counseling, pediatrics, and clinical genetics), and CRB parents with invaluable perspectives, lived experiences, and cultural insights, who collaborate on the research design, conduct, and materials of the ABGS study. Onboarding of new CRB members required education in genomic screening, along with its ethical, legal, and social implications (ELSI), to enable them to contribute informed perspectives regarding pediatric genomic screening. As this capacity-building exercise has expanded to the co-design of publicly accessible genetic health literacy resources via a community-academic partnership, members of the ABGS collaborative research team meet monthly via video conference and periodically in person to engage in an informal version of Community Engagement Studios [36] to discuss the research design, conduct, and materials of the ABGS study.

Initially, ABGS researchers curated capacity-building topics for the new CRB members to cover essential medical genetic concepts vital for building public health genetic literacy and facilitating informed decision-making regarding involvement in research or use of genetic health services (Table 1). Despite UNC research team members’ broad backgrounds in genetics research and clinical practice, we struggled to effectively convey crucial information to CRB members in an engaging and accessible manner. In early capacity-building sessions, CRB feedback played a pivotal role in shaping the trajectory of future resource development. The input from the CRB influenced decisions regarding content, length, delivery mode, readability, and ensured the resources resonated appropriately and effectively with their intended public audience.

**Table 1:** ABGS Genetics & Genomics Education Modules.

| Module Number | Title | Main Topics |
| --- | --- | --- |
| 1 | DNA – What is it and what does it do? | • What DNA and genes are<br>• How genes affect traits within families<br>• Connections between genes and disease |
| 2 | How Traits are Inherited | • Similarities and differences between each person’s DNA<br>• Genetic inheritance patterns<br>• Disease risk inheritance throughout families |
| 3 | Your Genes and Your Health | • How gene variants can affect the body’s function<br>• Examples of different types of gene changes (point mutation/deletions/missense)<br>• How gene variants can have different implications for health (variable expressivity/penetrance) |
| 4 | Genetic Testing and Screening | • Different purposes of genetic testing and screening<br>• Examples of when providers would perform genetic testing versus genetic screening<br>• How results for testing and screening differ and the process for returning results to patients |
| 5 | Safeguards in Genetic & Genomic Research | • Types of information used in genetic and genomic research<br>• Overview of prior harms in biomedical research that led to changes in research participant protections<br>• Informed consent and current safeguards protecting human research participants |
| 6 | Possible Risks and Benefits of Genetic Screening | • Examples of how genetic screening can affect public health<br>• Multilevel benefits and risks of genetic screening<br>• Informed consent and decision-making |
| 7 | All About the Age-Based Genomic Screening Program | • Description of ABGS approach<br>• Implementation of the ABGS pilot study<br>• Example conditions included in screen and what a participating family can expect during the screening process |

#### Evaluating Evidence, Needs, and Demand for Public Health Genetic Literacy

The increasing accessibility of genomic technology and information across clinical, commercial, media, and social media platforms has coincided with a surge in misinformation and gaps in genetic and genomic understanding. Consequently, there is a pressing need for resources to bridge these gaps outside specialized genetics environments, especially considering that individuals possess varying levels of literacy. Kaphingst et al. [29] distinguish between genetic knowledge and genetic literacy, highlighting that genetic literacy not only involves acquiring adequate knowledge of genetics and genomics but also entails the ability to effectively communicate and apply this knowledge in personal, social, and professional settings [37]. They underscore the importance of future empirical research in identifying the essential domains of genetics - and genomics-related skills and knowledge for the dissemination and utilization of genomic information in both public health and clinical settings, as well as understanding the relationship between these domains and the application of genetic knowledge [38,39]. We have adopted the Kaphingst et al. definition of genetic literacy from

During almost 20 planned or invited engagement and education events in NC, members of the ABGS collaborative research team actively participated to raise awareness of genetic and genomic among the public. ABGS participation aimed not only to educate but also to engage with community members, eliciting their thoughts, ideas, and concerns regarding genetic and genomic research. Through these interactions, specific community priorities have emerged prominently, guiding the development of educational resources tailored to address these urgent concerns. Community priorities included the demand for transparent information regarding the purpose and procedures of genetic and genomic research and health services, as well as questions and concerns about data storage, privacy, access rights, and the potential risks, benefits, and relevance to themselves, their families, and their communities. Additionally, community members have raised questions about whether genetic screening may reveal conditions not covered by health insurance or federal health programs, despite the availability of preventive or management strategies. Overall, community members persist in their desire to expand their understanding about genetic and genomic information, a factor known to increase recruitment of Black and African American participants in genomic research [40,41].

#### Prioritizing Equity in Context and Capacity for Change

Historical events of abuse and exploitation perpetuated by researchers from academic and medical institutions against racialized minorities, as well as marginalized, lower-resourced, and disabled individuals and communities in the U.S. has led to enduring mistrust of biomedical research and the healthcare system among these populations [21,42]. Furthermore, ongoing health discrimination and disparity, such as insufficient communication and exclusion from community research and developed outcomes, exacerbate this mistrust and discourage participation in genetic and genomic research, despite its potential to mitigate health inequities if implemented with representation from all populations [43–45]. Nevertheless, amidst these persistent barriers, many members of underserved communities maintain interest and an open-mindedness toward genetic and genomic research and public health interventions, driven by the prospect of disease prevention and an altruistic regard for future generations [25,46,47]. To resolve this tension, additional research into the best practices for developing and disseminating educational resources on genetics and genomics with particular emphasis on empowering and informing marginalized and harmed communities is necessary[48].

Current research emphasizes the communication gap that exists between underrepresented communities, increasingly targeted for participation in genomics research, and presumably well-intentioned yet culturally distant researchers [4,20,49]. These studies reveal a significant mismatch between study materials, recruitment strategies, and incentives, and the specific cultural contexts they are employed in, often resulting in a comprehension gap between researchers and the core values of the communities they aim to engage. Recognizing these discrepancies, it becomes necessary to adhere to the principles of communication science, and tailor engagement and recruitment approaches to align with the cultural nuances significant to the target communities. Fortunately, there is growing adoption of such practices, supported by an increasing number of funders, such as the Patient-Centered Outcomes Research Institute and the National Human Genetics Research Institute. The organizations are actively funding academic-community partnerships aimed at pursuing these inclusive practices. The D4DS paradigm offers valuable insights to optimize research products and innovations co-designed through these partnerships, ensuring broad reach and feasibility from their inception.

### Phase 2: Design

During the D4DS Design phase, collaborative efforts focus on developing research products and formulating active dissemination and sustainability plans that align seamlessly with the intended context. F2C objectives and outcomes aim to ensure congruence with the established needs, demands, and capacity for change within the community or population represented by the partners. Incorporating active dissemination and sustainability plans within the Design phase is crucial, as they require customization to messaging, packaging, and distribution channels that resonate effectively with the intended audience and setting. In the following subsections, we outline our current and ongoing efforts, including co-designing the Genetics and Genomics Educational Modules, preliminary dissemination of the modules within communities to establish bidirectional communication and receive feedback, implementing a feasibility pilot, and outlining future plans for sustainability.

#### Co-Designing Genetics and Genomics Educational Modules

Insights from early module drafts utilized in community health events broadened the scope of resources being co-developed by the ABGS collaborative research team into seven comprehensive Genetics and Genomics Educational Modules (“the modules”) (Table 1). Initially conceived of and developed as presentation slide decks, they serve as dynamic storyboards, providing a flexible framework for the development of various media formats. Each module follows a structured format comprising an introductory message, a main body of educational content, and a set of ‘takeaway concepts’, with an average presentation duration of under 15 minutes. To ensure the accessibility of the content, the modules are crafted in plain language, with readability tailored to a 5th to 8th-grade level. Consistent language and illustrations are used across all modules to maintain coherent messaging throughout the sequence, although they can be presented either sequentially or as standalone learning objects. Customized scripts are designed to facilitate both independent and collaborative presentations.

The first four modules, completed at the time of publication, delve into the scientific principles of genomic medicine. The subsequent duo of modules, currently at various stages of review and development, explore ethical, legal, social, and technological dimensions of genetic and genomic screening and research. These modules address complex topics such as historical instances of unethical biomedical research and the contemporary research safeguards developed in response. The planned Module 7 will further build upon the content covered in the previous modules by introducing the ABGS study itself.

Initially, the co-development process followed a linear path (Figure 2A) with a small writing team comprising UNC researchers creating a basic draft of the module. Then this team gathered and incorporated feedback from the CRB through templated Google forms and video conference discussions, with a focus on assessing the appropriateness, accessibility, relevance, and clarity of the genetic concepts and images. This feedback loop was then replicated with the UNC research team, who provided input regarding the accuracy and depth of the content. ABGS researchers within the writing team then integrated this feedback to refine the modules, ensuring better alignment with the target audience’s needs.

**Figure 2:**
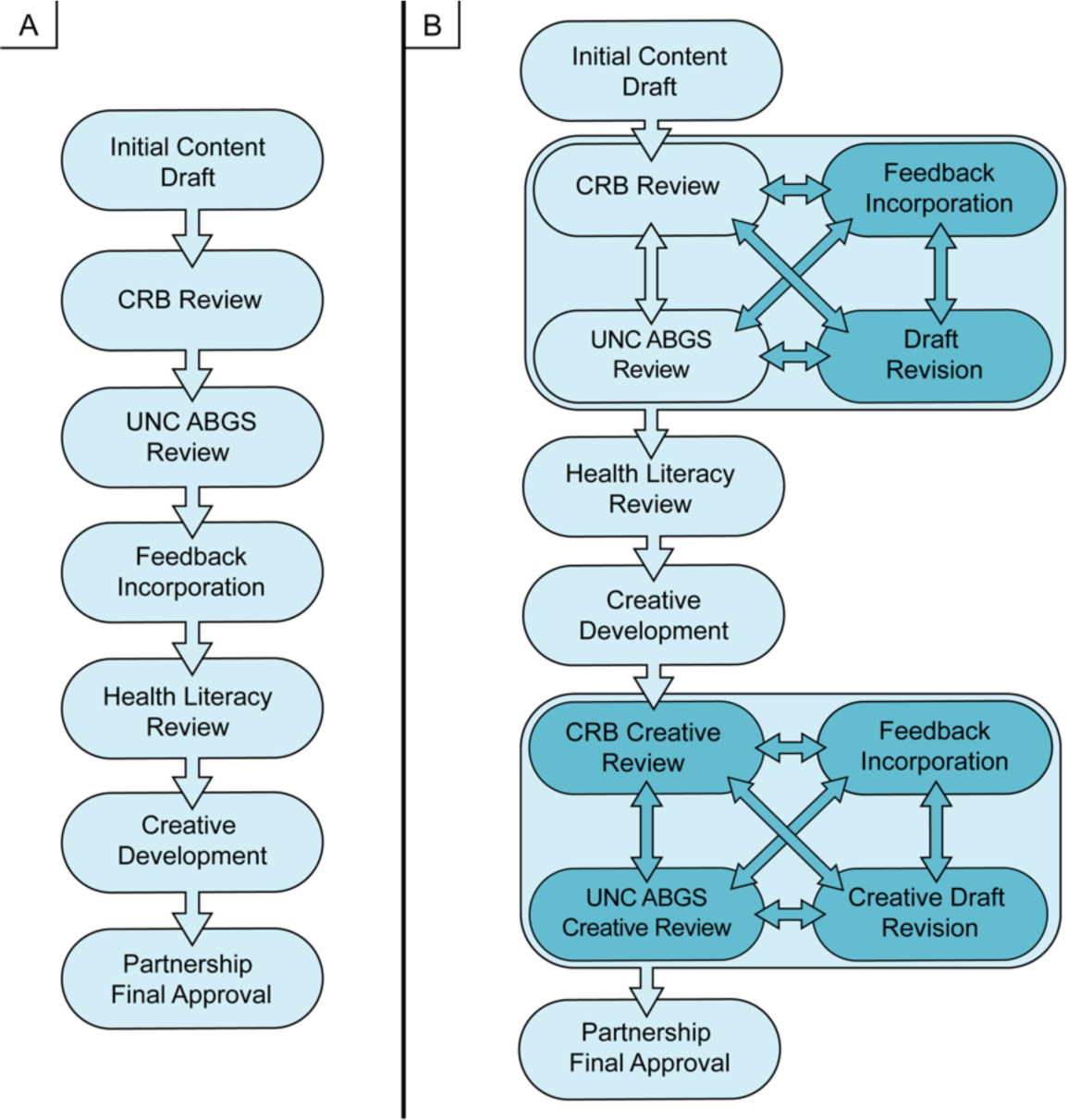
(A) Initially, a linear process was employed for the co-design of the modules. (B) An expanded approach to module creation was later adopted, entailing iterative cycles of reviews among the subgroups of the ABGS collaborative research team. This iterative approach was implemented to ensure the incorporation of diverse perspectives and insights, enriching the developmental process of the modules.

Following this, a health literacy librarian from UNC’s Health Sciences Library assessed readability using the metrics of Flesch-Kincaid Reading Grade Level and SMOG (Simple Measure of Gobbledygook)[50], identified areas of unnecessary complexity, and suggested plain language terms for broader accessibility. Following the literacy review, the ABGS researchers ensured the module content retained its scientific veracity. To address the dilemma of balancing readability with scientific precision, the ABGS collaborative research team began co-developing an accompanying plain language glossary of essential genetic terms as a supplement to the modules.

Ultimately, the challenges encountered during the co-creation process, balancing visual and scientific accuracy and consistency, while ensuring broad accessibility, prompted the team to transition from a linear to an iterative design approach. The diverse composition of the CRB led to a spectrum of recommendations, at times conflicting with each other, or with the research team’s understanding of patient education practices. Although this adjustment added complexity to tracking and incorporating feedback, we believe it fostered a more inclusive approach where all perspectives were equally heard and considered.

The new iterative design process utilized in module development involved multiple rounds of review among researchers and between UNC researchers and the CRB at various stages (Figure 2B). Following each iteration, the module underwent refinement, resulting in a more polished version [51] that advanced to the next stage. Given the added complexity of the review process, we established documentation practices to accurately manage feedback and suggestions arising from each review cycle. During deliberations on the suggestions, ABGS researchers documented the rationale behind each decision, sharing them with the CRB. Revisions underwent a review process for group consensus, often resulting in additional changes aimed at strengthening the connection between the content and the intended audience. The number of review rounds needed to finalize each module varied, influenced by factors such as topic complexity and the extent of feedback required. In cases where consensus could not be reached, executive decisions were made by the ABGS principal investigators to avoid collaborative stalemates. Following these rigorous iterative cycles, the modules were translated into comics and custom-illustrated slides accompanied by scripts for public distribution.

#### Engaging Communities in Dissemination and Education

We prioritized accessibility by integrating user-friendly graphics alongside the content. Farinella showed that illustration, particularly in comic-style format, is highly effective in medical and science education [51]. This approach facilitates understanding of complex concepts by presenting engaging narratives with playful characters and visual metaphors. We posit that incorporating illustrations could enhance the accessibility of complex genetic topics, especially for individuals with limited prior knowledge in the field. To accomplish this goal, the ABGS collaborative research team collaborated with a professional science illustrator to craft vibrant comic-style graphics. These illustrations feature an iconic anthropomorphic narrator, “Gene”, who simplifies key genetics educational concepts for a lay audience through straightforward visuals, analogies, and relatable examples. These graphical and narrative elements are consistently applied across all module storyboards to maintain coherence. During development, we utilized these illustrations in comic books that encapsulate the content of each module. These comics serve to reinforce the concepts presented in the slides or offer an independent modality for conveying ideas about genetics and genomics. Additionally, the comics underwent iterative reviews by the ABGS collaborative research team to ensure alignment with the drafted module content in the storyboards and to secure full team approval before subsequent dissemination.

The module comics, available in both full-color editions and black-and-white coloring book versions, are distributed at various community events across North Carolina. These events, such as health fairs, STEM expos, workshops at high schools, and Heritage Month celebrations, serve as platforms for engaging with diverse communities and fostering awareness and interest in genomic medicine, by engaging the public in discussions with representatives of the ABGS project (including both researchers and CRB members). By encouraging productive interactions prior to recruiting parents for the ABGS pilot study, our objective is to establish an open exchange of information between researchers and the community. This lays the groundwork for fostering relationships and bidirectional communication with stakeholders in potential implementation areas. The initial feedback garnered from individual community members at these events has shown a great deal of enthusiasm for the design and content of the comics. By connecting with members of diverse communities through accessible educational content, we can spark discussions about genetics and screening perceptions, thereby acquiring valuable insights into the learning needs and preferences of different community members (Table 2).

**Table 2:** Examples of ABGS Community Outreach.

| Event | Audience | Mode | Medium | Modules (#) |
| --- | --- | --- | --- | --- |
| Lunch & Learn sessions | Community members and champions | Virtual | Slide presentation | 1-5 |
| School Presentations | Parents, teachers, administrators | Virtual/<br>In-person | Comics, slide presentation, videos, infographics, interactive electronic content | 3, 4, & 6 |
| ABGS Focus Group | Parents & guardians | In-person | Slide presentation, videos, infographics | 3, 4, & 5 |
| Community Outreach Events | General public | In-person | Comics, infographics, interactive electronic content | 1-7 |
| Onboarding | New ABGS-CRB partnership members | Virtual/<br>In-person | Slide presentation | 1-7 |

The storyboards, the initially created slide presentations, offer a flexible format for the development of additional future media types such as brochures, animated videos, interactive web content, and infographics. This approach allows us to customize our outreach efforts to suit diverse audiences. By leveraging the same storyboard for multiple media types, we can streamline the creation process and ensure consistency in messaging. To better understand community preferences regarding messaging, packaging, and distribution channels, the ABGS team incorporated two survey questions into a poster board displayed at community engagement events, allowing attendees to share their preferences independently. Participants indicated their interest level in learning about genetic screening and their preferred method of learning this information through “dot voting”, marking their choices by placing sticky dots on relevant categories. The first question gauged interest in hypothetical genetic screening in children for actionable genetic conditions, offering response options ranging from “very interested” to “not at all interested.” The second question prompted participants to select their top three preferred methods for learning about genetic screening for children from the choices: attending live educational events, watching videos, visiting websites, playing app-based games, engaging with social media content, reading pamphlets, consulting with a child’s doctor, and others.

Preliminary results from over 500 attendees across four community events held within North Carolina’s Research Triangle area in 2023 and 2024 indicate that most respondents are at least “somewhat interested” in genomic screening. From this ad hoc approach for obtaining information about preferences, the top three methods for learning more about genomic screening are visiting a website, watching a video, and consulting with the child’s doctor. This feedback underscores the importance of online resources and healthcare provider guidance in disseminating information about genomic screening to the public, and suggests that once complete, we should prioritize adapting the modules to video.

#### Mixed-Methods Feasibility Study

Building upon the preliminary feedback and interactions with community members, which indicated learner preferences for genetic and genomic educational materials, we devised an explanatory mixed-methods feasibility study. This study aims to investigate whether the modules can augment genetic health literacy and subsequently bolster the likelihood of research participation. Simultaneously, as we introduce the modules to facilitate meaningful interactions with the public at community events, we are examining the impact of the educational content and delivery methods on event attendees. Consented participants are randomly assigned to one of four groups, each exposed to different educational material and delivery methods. They are then asked to view this material and complete pre- and post-surveys measuring genomic knowledge and willingness to participate in genomic research. Additionally, a diverse subset of survey respondents will undergo interviews to explore their perceptions and preferences regarding learning about and engaging in genomic research.

Conceptual frameworks and models from D&I science are increasing being adapted for the application of an equity lens to the assessment of context and implementation [52,53]. To this end, we developed a semi-structured interview guide based on multi-level contextual domains from PRISM [33] with modifications by Fort et al. [54] to emphasize health equity. We also utilized the health equity implementation framework (HEIF), which was specifically developed for assessing both implementation and health equity determinants [55].

Through this pilot study, our goal is to gather insights into preferred learning modalities and sources of information in genetic and genomic medicine, thereby assessing determinants likely to influence dissemination, adoption, and sustainability. Ultimately, we seek to uncover issues with research participation that may not have been addressed during engagement events and to gain a contextual understanding of individuals’ perceptions of the modules and delivery methods. By comparing and integrating data from quantitative and qualitative sources, we anticipate gaining a richer and more comprehensive understanding of how genomic knowledge and community members’ perceptions impact their decision-making about participating in genomic research. This understanding will guide further refinement of the modules to prepare for a larger rollout to various communities and invested users.

#### Adapting Module Content for Future Sustainability

The slideshow presentations, full-color comics, and coloring book versions of the modules are licensed under a Creative Commons license “CC BY” (Creative Commons, n.d.), accessible at https://creativecommons.org/licenses/by/4.0/. Once finalized, they will be readily available for download on the “Parent &Community Engagement” page of the ABGS website (www.med.unc.edu/genetics/abgs). Early drafts of the modules serve as tools to foster active and reciprocal networks with local community representatives. These individuals, intended champions of the modules’ use and dissemination, are pivotal partners possessing significant insight, influence, and access within their respective communities. Establishing and nurturing such trusting relationships is fundamental for the ABGS project’s ability to effectively respond to the needs of target communities, including addressing health equity priorities and creating culturally appropriate, feasible, and acceptable content. These connections play a crucial role in identifying effective communication channels and delivery methods [56].

### Phase 3: Dissemination

During the D4DS Dissemination phase, the research product developed in the preceding phase is actively shared through accessible and culturally appropriate methods and communication channels. In this phase, our priorities will include expanding and enhancing community partnerships through capacity-building activities, utilizing optimal content distribution methods in diverse communities, translating content into Spanish to promote equity, and engaging with clinical providers and other potential adopters and champions to integrate the use of the modules into genetics and genomics research and practice.

#### Building Equitable Access Through Language Translation

To expand our outreach efforts, we have established a bilingual ABGS Spanish translation team, comprised of a genetic counselor, a graduate student, and a research assistant. The primary goal of this team is to ensure that all modules are accessible in both English and Spanish. We selected Spanish due to its prevalence as the second most spoken language in North Carolina, with 7.9% of the population exclusively using Spanish, and over 1.1 million people in the state identifying as Hispanic/Latino [57]. To guarantee accuracy and accessibility, our Spanish translation team, along with several bilingual members of the CRB, engage in an iterative process of translation, review, and discussion. This approach mirrors the iterative review process used for the modules, aiming to maintain the fidelity of translated materials while prioritizing readability, accessibility, and comprehensibility for the Spanish-speaking audience.

The bilingual members of the ABGS collaborative research team have facilitated our participation in community outreach at Spanish cultural events. Our early outreach with Spanish-speaking populations has shown considerable interest in genomic screening for children. We have observed that delivering information in a familiar language fosters inclusivity and greatly enhances enthusiasm for engaging with the material. Engaging with community members in multiple languages is crucial, and our commitment to providing unbiased and fact-based information serves as a powerful tool in fostering genuine connections and dispelling prevalent misinformation.

#### Disseminating Modules through Community Outreach

Critical for ensuring sustainability, substantial capacity-building activities will be required to establish successful community partnerships for disseminating the modules within communities of interest. The conceptualization and design of these modules were significantly influenced by prior engagement and awareness efforts, as well as data collection with various partners advocating for health equity among underserved groups including refugee/immigrant families, Latino/a and Hispanic, Asian Americans and Pacific Islanders, and Black communities, and rural communities in eastern and western North Carolina. Tailored strategies resulting from these engagement activities will aim to promote health equity by positively influencing dissemination and process outcomes, fostering trust, connection, interest, and uptake (Table 3). We will leverage the results of the mixed methods study to further refine the modules before disseminating them to a wider audience through collaborations with public adopters and champions.

**Table 3:**
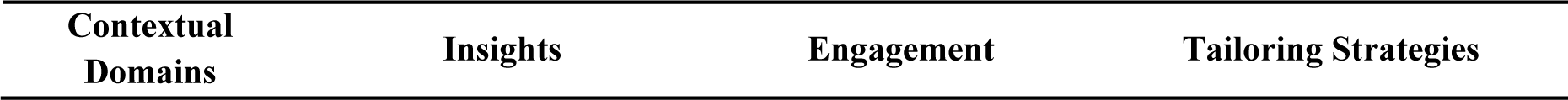
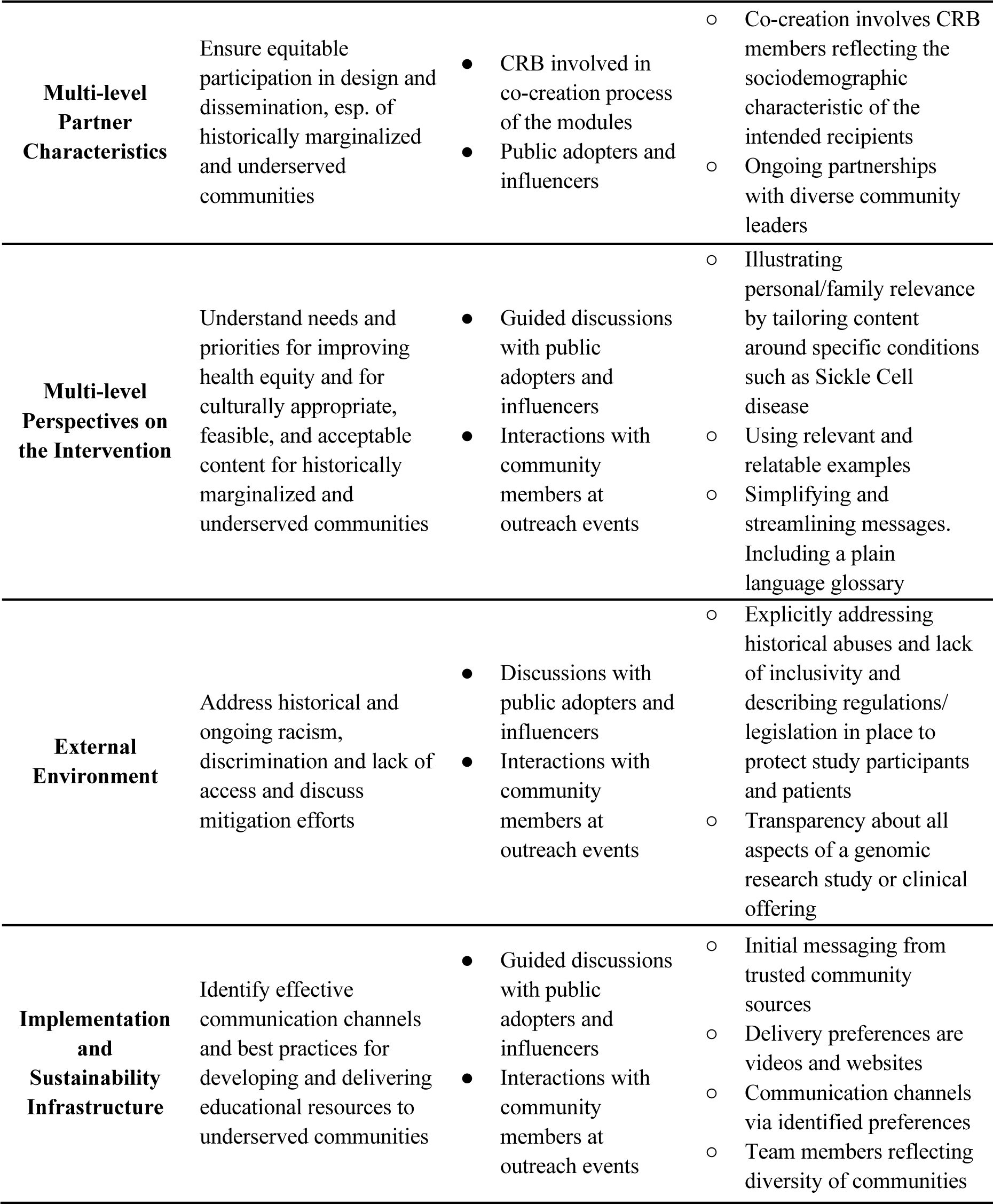
PRISM contextual factors, partners, insights, and impacts.

| Contextual Domains | Insights | Engagement | Tailoring Strategies |
| --- | --- | --- | --- |
| **Multi-level Partner Characteristics** | Ensure equitable participation in design and dissemination, esp. of historically marginalized and underserved communities | • CRB involved in co-creation process of the modules<br>• Public adopters and influencers | ○ Co-creation involves CRB members reflecting the sociodemographic characteristic of the intended recipients<br>○ Ongoing partnerships with diverse community leaders |
| **Multi-level Perspectives on the Intervention** | Understand needs and priorities for improving health equity and for culturally appropriate, feasible, and acceptable content for historically marginalized and underserved communities | • Guided discussions with public adopters and influencers<br>• Interactions with community members at outreach events | ○ Illustrating personal/family relevance by tailoring content around specific conditions such as Sickle Cell disease<br>○ Using relevant and relatable examples<br>○ Simplifying and streamlining messages. Including a plain language glossary |
| **External Environment** | Address historical and ongoing racism, discrimination and lack of access and discuss mitigation efforts | • Discussions with public adopters and influencers<br>• Interactions with community members at outreach events | ○ Explicitly addressing historical abuses and lack of inclusivity and describing regulations/legislation in place to protect study participants and patients<br>○ Transparency about all aspects of a genomic research study or clinical offering |
| **Implementation and Sustainability Infrastructure** | Identify effective communication channels and best practices for developing and delivering educational resources to underserved communities | • Guided discussions with public adopters and influencers<br>• Interactions with community members at outreach events | ○ Initial messaging from trusted community sources<br>○ Delivery preferences are videos and websites<br>○ Communication channels via identified preferences<br>○ Team members reflecting diversity of communities |

A multifaceted educational and awareness campaign will require a diverse array of communication methods tailored to the preferences of the communities involved. Visual aids, including illustrations and videos, play a crucial role in enhancing content engagement [35,58–60]. Utilizing popular platforms like Facebook, TikTok, and YouTube has proven effective [59,61] for reaching diverse audiences actively seeking health related resources and we plan to promote the modules via these outlets in the future [62]. However, there remains a significant need for research on best practices for designing educational resources aimed at increasing public interest and willingness to participate in genomic research, particularly within underserved and marginalized communities [63]. As we analyze the data from the mixed methods study, we will assess whether the specific content and/or method of delivery of educational materials used in participant engagement efforts influences willingness to participate in research.

Bilingual members of the CRB have suggested utilizing the translated modules, once completed, to educate and engage bilingual community health workers (CHW) as agents for disseminating and enhancing capacity through education and training. Leveraging CHWs, who often have deep ties within the communities they serve, can foster trust and acceptance of new initiatives. By involving CHWs in the dissemination and ongoing adaptation of modules, we can ensure that the content remains relevant and effective across diverse community contexts.

#### Connecting with Primary Care Providers

Ultimately, ABGS implementation will entail primary care providers (PCPs), including pediatricians, family medicine physicians, physician assistants and nurse practitioners, integrating genomic screening into routine well-child visits, and presenting information to parents in clear, concise language. This approach aims to enhance clinical utility and improve accessibility of genomic screening. Regular discussions between providers and parents, along with gradual introduction of the child to genomic concepts, are expected to foster genetic literacy and confidence in making informed decisions among older children and adults.

However, the widespread adoption of pediatric genomic screening in primary care settings will require addressing implementation challenges since this paradigm shift moves genomic screening from specialized settings to primary care. Research indicates that many PCPs feel inadequately prepared to discuss genetic offerings with patients [64–67]. We believe that the modules could serve as a valuable resource in this context, potentially assisting providers in navigating conversations about research or clinical offerings. While additional resources will be needed to support PCPs in obtaining consent and managing results, the modules offer a promising tool to help providers gain confidence in discussing genetic and genomic research, testing, and screening with their patients.

### Phase 4: Impact

During the D4DS Impact phase, the research product is evaluated to gauge its effects on both health and health equity, while also considering continual alignment with contextual factors and the necessity for adaptation. In this phase, we will use RE-AIM for multilevel evaluation of external validity over a variety of settings.

#### Assessing and Adapting the Modules for Equitable Impact

To ensure equitable impact, we iteratively assess RE-AIM outcomes to plan and organize the dissemination of the modules into communities, as well as to identify equity-enhancing approaches and address any unintended consequences of adaptations (Table 4). In addition to surveys and qualitative measures, we will utilize automated approaches such as YouTube Studio channel analytics (e.g., impression rates, unique viewers, click-through rates), Google Analytics metrics for the project website (e.g., session duration, pages per session, source of traffic), and patient administrative data (with consent) from clinic partners (e.g., gender, age, race, ancestry). Emphasizing continuous improvement over time is crucial for sustaining the impact of these modules. As we identify more opportunities for adoption and receive requests for adaptations, we will plan to remain responsive to the evolving needs of communities. An iterative process will enable us to refine the modules to better address the unique challenges and circumstances faced by different populations.

**Table 4:**
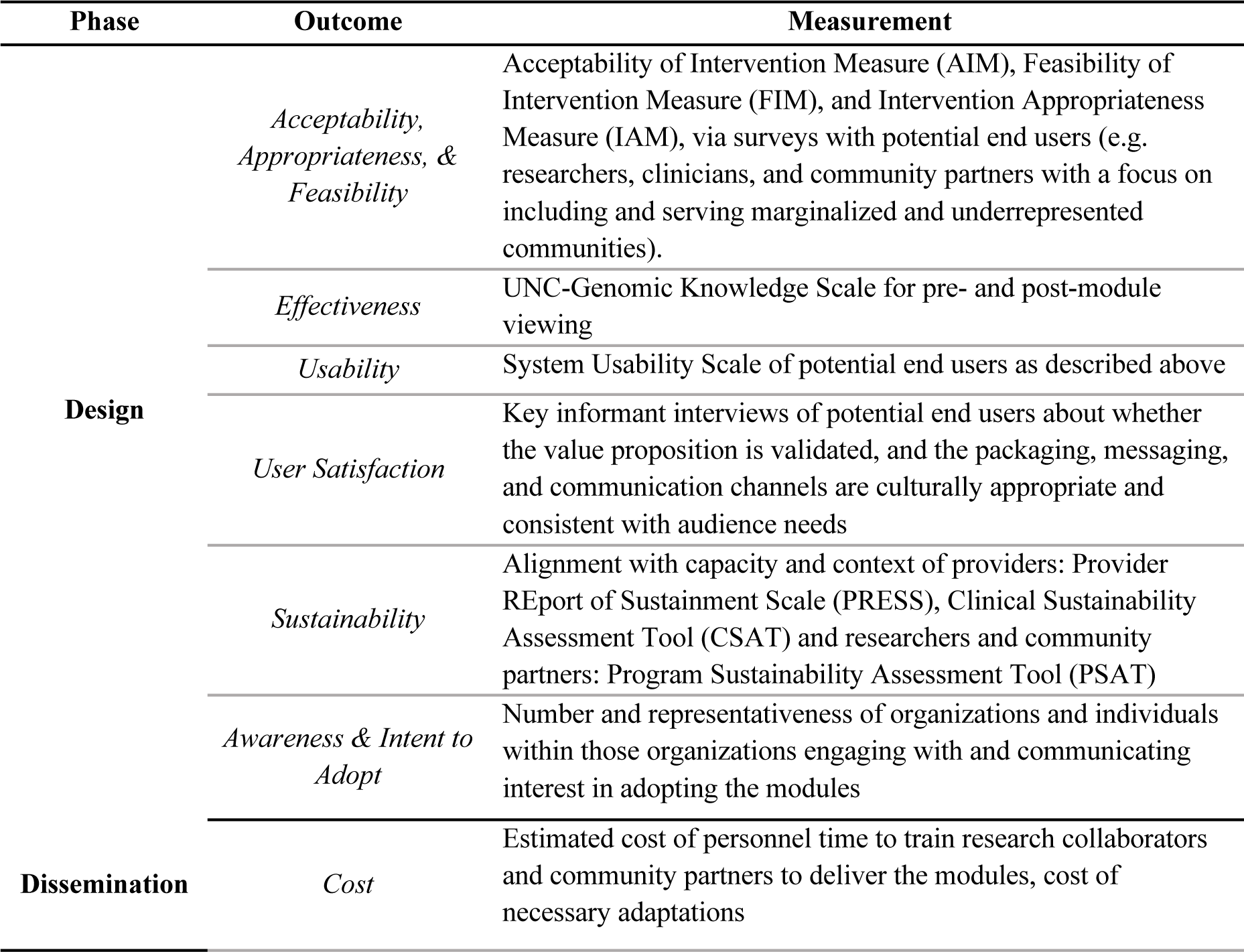
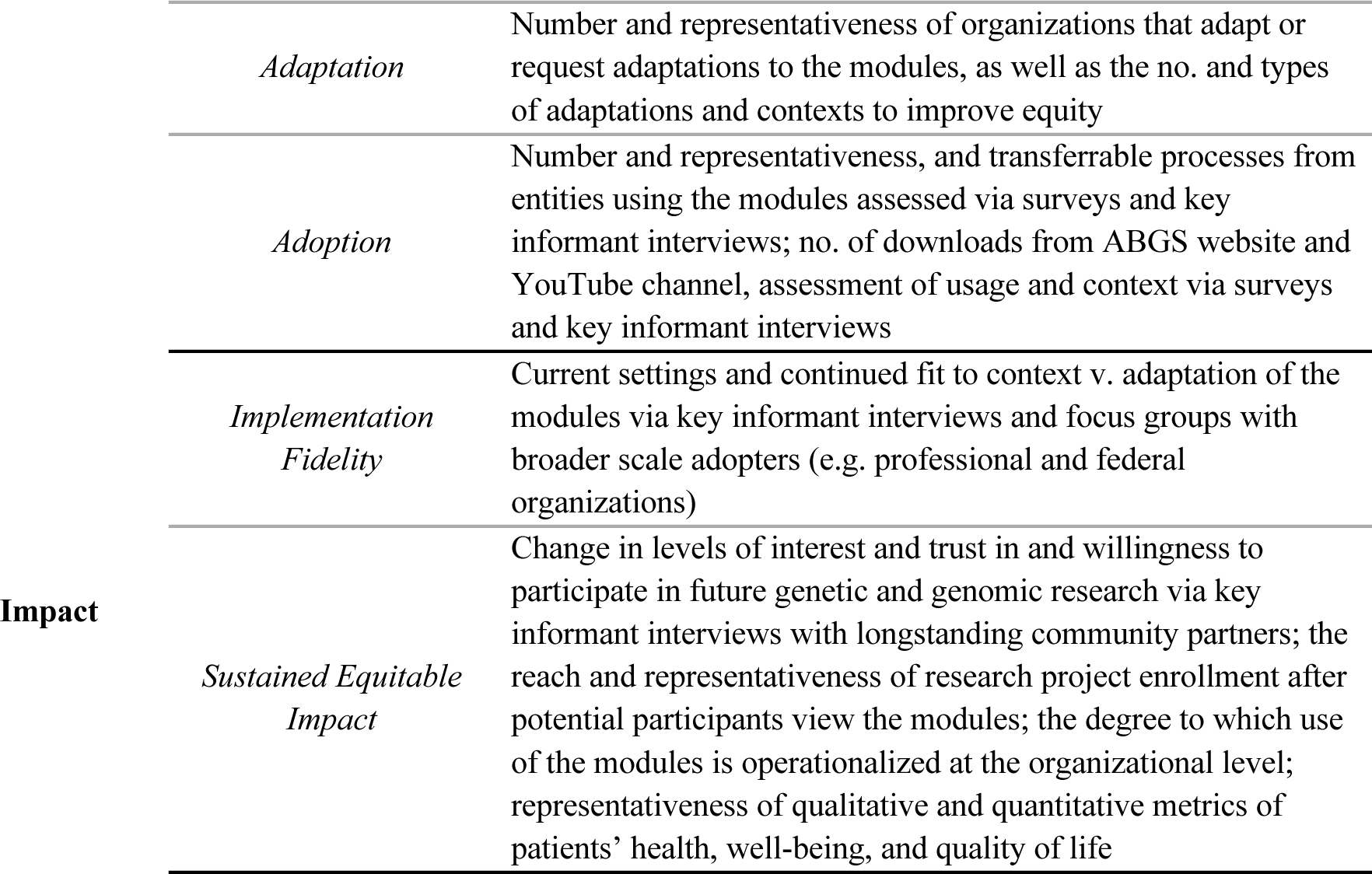
RE-AIM framework applied to the Module Development process.

## Discussion

Research investigating both professional and public perspectives on genomic screening in individuals of all ages indicates significant interest but also persistent apprehension regarding the implementation of such screening [66,68–71]. For parents considering genomic screening for their children, concerns range from anxiety over decisions concerning the disclosure of genetic information and data security to prioritizing immediate needs over elective genomic sequencing [69,72]. Additionally, worries about costs, future discrimination, and the psychological impact of discovering untreatable health conditions are common [70]. Effective and equitable integration of genomic screening into pediatric well-child care requires the establishment of trust with diverse community partners to discern sought-after information and determine optimal communication methods. Left unaddressed, these individual concerns could erode public trust in and impede the uptake of public health screening initiatives aimed at reducing health disparities.

The widespread deficiency in genetic health literacy among the public significantly impacts both clinical research outcomes and patient care. Furthermore, this gap is widened by various sociodemographic and other factors, resulting in a pronounced discrepancy in comprehension and communication between researchers and community members [2,20]. This discrepancy can particularly affect the recruitment and enrollment of underserved individuals into genetic and genomic research. To address this challenge, we have taken a proactive approach to better understand community perspectives on enhancing public health genetic literacy before attempting to recruit or enroll community members for ABGS. In line with the D4DS paradigm’s emphasis on co-designing user-friendly bilingual educational resources, our ABGS collaborative research team worked closely with a science illustrator and a graphic artist. Together, we iteratively crafted modules that integrated feedback from both community members and experts, ensuring information is presented in an engaging and accessible manner.

The community-academic partnership forged by the ABGS collaborative research team has significantly enhanced the value of the modules. Drawing on diverse perspectives, we worked to ensure that our materials resonated with the communities we serve and effectively conveyed crucial information about genetics. In addition to module development, our team actively engaged with the public to gain insight into the needs of underserved communities. During these engagements, community leaders emphasized the importance of reliable information to facilitate equitable access to healthcare advancements, including genomic screening. They expressed a need for educational materials that address topics such as family history discussions, modes of genetic inheritance, and the importance of sharing genetic testing and screening results with close blood relatives. Furthermore, interactions with underserved communities underscored the need for educational resources presented in lay language. Such materials not only promote genetic literacy but also foster trust between these communities and the medical establishment. They play a crucial role in facilitating informed decision-making and ensuring equitable participation in research.

Through our outreach efforts to community members, we also became aware of the need to carefully understand the public perceptions about the effectiveness of genetic and genomic testing and screening in different populations. Many people are aware that most genetic research has been done in individuals of European ancestry and that research is lagging for other ancestral backgrounds. This is especially true of research in common complex diseases, where studies require thousands or tens of thousands of cases and controls, and the genetic architecture of disease risk may vary across different ancestral populations. However, due to the nature of rare disease research, disparities play a more nuanced role in the effectiveness of genetic testing for Mendelian/monogenic conditions. It can therefore be difficult to disentangle the perceived and actual impact of disparities in research participation from the potential benefits and limitations of genomic screening for each individual.

Greater participation in genetic research and data sharing will enrich the knowledgebase about disease-causing variants, thereby promoting the virtuous cycle of improving equity of genetic testing outcomes. This nuanced understanding is pivotal, serving as a crucial equity marker for genetic health services, and forming the basis for a compelling value proposition aimed at encouraging participation in genetic research and data sharing among these communities. Furthermore, a significant health equity concern lies in the restricted access to cutting-edge research and potential therapies within racially minoritized populations. Clearly addressing these issues in educational resources not only fosters inclusivity but also strengthens the incentives for engagement in genetic and genomic research within these communities.

We encountered several barriers during the project. Coordinating input from researchers and clinicians during the module development proved demanding, as did the limited availability of resources to conduct outreach. Balancing feedback from various invested users, including researchers, clinicians, and CRB members required careful handling to prevent conflicts within the ABGS collaborative research team and ensure that all perspectives were thoroughly considered. To address this, we developed a multilevel pathway to ensure all feedback was accurately documented. Disseminating the modules at outreach events has presented constraints including insufficient personnel or material resources to attend events. To overcome this, we will focus efforts on areas of highest need for populations underserved in genetics, collaborating with community partner organizations to explore more intimate settings to deploy the modules, such as Lunch & Learns and listening sessions rather than broader community-wide events, such as fairs or street festivals.

Our approach is centered on collaborative development and eventual widespread utilization of the modules. We prioritize adaptability and flexibility to incorporate feedback from a diverse array of community partners. This iterative strategy allows us to continually improve the modules, making them more relevant and effective, in line with the principles of D4DS methodology and the F2C framework. Thus, our collaborative efforts extend beyond the collaboration between ABGS researchers and CRB members to create the educational modules. Our efforts also encompass broader community outreach, including presentations and dissemination to local public-school classrooms, clinical providers, community members and connectors, as well as research collaborators, with the aim of eliciting responses and engagement from diverse potential end users. While our exploration of these concepts is centered on participation in genomic research, we advocate for the wider adoption of this methodology by researchers to foster more equitable relationships with all communities.

## Conclusion

The effective translation of genomic discoveries into equitable public health contexts hinges on understanding and addressing genetic literacy. This will require further research to define genetic literacy, establish guidelines for proficiency, and develop interventions tailored to individuals with varying levels of overall literacy. Drawing from related fields such as health communication, health services research, and dissemination and implementation science, interventions can be designed to enhance communication and information dissemination. User-centered design approaches, including co-creation with community members, are essential for developing culturally and linguistically responsive genetic literacy interventions and dissemination strategies. Collaboration among researchers, practitioners, and community members is vital for advancing public health genetic literacy and optimizing the use of genomic information in research, healthcare, and society at large.

## Statements

## Acknowledgements

We would like to thank Terri Ottosen, UNC health literacy librarian, and Sharisse Jimenez, a core member of the Spanish translation team, as well as Rachel Phillips for assistance with grammatical edits. We would also like to acknowledge with gratitude many community members who have freely shared their valuable insights to inform the co-development of the modules.

## Statement of Ethics

Please address the following aspects in your Statement of Ethics.

### Study approval statement

The future evaluation of the module through a study protocol has been reviewed and been given an exemption by the UNC Institutional Research Board under study number 23-2772.

### Consent to participate statement

In future evaluation of the module content with mixed-methods studies, any study participants will be required to give written consent for any surveys or interviews conducted.

## Author Contributions

Grace Byfield, Kimberly Foss, Sabrina Powell, Daniel Torres, Ann Katherine Major Foreman, Jahnelle Jackson, Grace Leon-Lozano, Juhi Salunke, Margaret Waltz, Julianne O’Daniel, Bradford Powell, Samantha Schilling, Megan C. Roberts, Stefanija Giric, Elizabeth Branch, Jonathan Berg, Laura Milko, Thomas Owens, Ken Ray, Carla Robinson, Lennin Caro, Erin Song, Andreas Orphanides, Jonathan Shaw, and Nicole Shaw contributed to the conception and design of the manuscript and modules. Grace Byfield, Jahnelle Jackson, Grace Leon-Lozano, Laura Milko, Sabrina Powell, Daniel Torres, and Juhi Salunke wrote the first draft of the manuscript. All authors contributed to the development of the Genetics and Genomic Modules and the manuscript revision and read and approved the submitted version.

## Data Availability Statement

The original contributions presented in the study are included in the article/supplementary material, further inquiries can be directed to the corresponding author.

